# SARS-Cov-2 prevalence, transmission, health-related outcomes and control strategies in homeless shelters: systematic review and meta-analysis

**DOI:** 10.1101/2021.01.14.21249851

**Authors:** Amir Mohsenpour, Kayvan Bozorgmehr, Sven Rohleder, Jan Stratil, Diogo Costa

**Affiliations:** Department of Population Medicine and Health Services Research, School of Public Health, Bielefeld University, Germany; Section for Health Equity Studies and Migration, Department of General Practice and Health Services Research, Heidelberg University Hospital, Germany; Institute for Medical Informatics, Biometry and Epidemiology, Ludwig-Maximilians-University Munich, Germany

**Keywords:** people experiencing homelessness, homeless shelters, SARS-Cov-2, Covid-19, systematic review, meta-analysis

## Abstract

**Background:** People experiencing homelessness (PEH) may be at particular risk for COVID19. We synthesised the evidence on SARS-Cov-2 infection, transmission, outcomes of disease, effects of non-pharmaceutical interventions (NPI), and the effectiveness of targeted strategies for infection prevention and control (IPC).

**Methods:** Systematic review of articles, reports and grey-literature indexed in electronic databases (EMBASE, WHO-Covid19, Web of Science), pre-print repositories, institutional websites, and handsearching. Empirical papers of any study design addressing Covid-19 in PEH or homeless shelters’ staff in English were included. (PROSPERO 2020 CRD42020187033)

**Findings:** Of 194 publications, 13 studies were included (two modelling, ten observational and one qualitative study). All were conducted in high-income countries. Random-effect meta-analysis of prevalence estimates yields a baseline SARS-Cov-2 prevalence of 2·14% (95% Confidence-Interval, 95%CI=1·02-3·27) in PEH and 1·72 % (95%CI=0·31-3·12) in staff. In outbreaks, the pooled prevalence increases to 29·49% (95%CI=16·44-29·55) in PEH and 15·18% (95%CI=8·95-21·42) in staff. Main IPC strategies were universal and rapid testing, expansion of non-congregate housing support, and individual measures in shelters (bed spacing, limited staff rotation).

**Interpretation:** Up to 30% PEH and 17% staff are infected during outbreaks of SARS-Cov-2 in homeless shelters. Most studies were conducted in the USA. No studies were found on health-related outcomes or health effects of NPI. An overview and evaluation of IPC strategies for PEH, including a better understanding of disease transmission, and reliable data on PEH within Covid-19 notification systems is needed. Qualitative studies may serve to voice PEH experiences and guide future evaluations and IPC strategies.

**Funding:** No source of funding.

**Panel 1: Research in context:** *Evidence before this study:* People experiencing homelessness (PEH) are at increased risk of infectious, chronic, and mental health adverse conditions. Due to the risk of transmission in shared accommodations, PEH may be particularly vulnerable to SARS-Cov-2 infection and worse clinical outcomes. Non-pharmaceutical interventions (NPIs) taken to mitigate the SARS-Cov-2 outbreak may have further aggravated health and social conditions. However, there is no evidence synthesis on the SARS-Cov-2 epidemiology among PEH, the correspondent clinical and other health-related outcomes as well as health effects of NPIs on these groups.

**Added value of this study** We reviewed and synthesized existent evidence on the risk of infection and transmission, risk of severe course of disease, effect of NPIs on health outcomes and the effectiveness of implemented measures to avert risks and negative outcomes among PEH. Results of the identified studies suggest that both PEH and shelter staff are at high risk of SARS-Cov-2 infection, especially in case of a local outbreak. Due to the low prevalence of symptoms at the time of a positive SARS-Cov-2 test among PEH, symptom screening alone may not be efficient to control outbreaks. Instead, universal and rapid testing conjugated with expansion of non-congregate housing support, and individual measures in shelters, are discussed as sensible strategies.

**Implications of all the available evidence** A comprehensive overview of NPIs and shelter strategies targeting PEH and evaluation of their effectiveness and unintended health consequences is needed. Qualitative research considering living realities of PEH can facilitate understanding of their specific needs during the pandemic.

## Introduction

Homelessness is associated with mental health problems^1,2^, infectious diseases^3^, cardiovascular and respiratory diseases, and several long-term conditions (e.g., asthma, COPD, epilepsy)^4^. People experiencing homelessness (PEH) are a diverse group categorized according to their living situation: (a) Rooflessness, people without shelter sleeping rough in the streets or in public spaces, (b) houselessness, with accommodation of temporary nature, (c) living in insecure housing, shaped by threats of insecure tenancy, eviction or domestic violence, and (d) living in inadequate housing, e.g. in caravans on illegal campsites or in extreme overcrowding^5,6^.

The policy measures taken to mitigate the spread of the SARS-Cov-2 pandemic, such as physical distancing and national lockdowns, may have aggravated their health and social conditions, adding to their already marginalised situation^7–9^.

Typical means of survival in daily life of PEH were disrupted, e.g. food banks and other basic aid facilities were shut-down, temporary housing projects were halted^8^. It is unclear how PEH dealt with the generalised closure of institutions, restrictions in movements and public transports, or how they perceived and adhered to the measures imposed, such as the use of nose and mouth protection.

PEH are vulnerable to SARS-Cov-2 infection due to the risk of transmission in shared accommodations, comorbidities and lower immune-response because of poor nutrition and food insecurity. Yet, evidence is scarce about the risk of infection among PEH, their differential susceptibility and health outcomes observed during the pandemic, to inform the design of infection protection and control (IPC) strategies.

We aimed to synthesize the evidence on the risk of infection and transmission, risk of severe course of disease, effect of non-pharmaceutical interventions (NPI) on health outcomes and IPC strategies to avert risks and negative outcomes among PEH.

## Methods

### Search strategy and selection criteria

The review protocol was registered in PROSPERO(PROSPERO registration 2020 CRD42020187033)^10^. The recommendations by the taskforce on guidelines for systematic reviews in health promotion and public health^11^ were followed. A systematic search of scientific databases was conducted in EMBASE, the WHO Covid19 database, and Web of Science Core Collection. The research questions are presented in Box 1. The search strategy details are presented in supplementary material 1.

#### Panel 2 research questions

The systematic review was guided by five specific research questions:

1. What is the prevalence or incidence of infection with SARS-CoV-2 in homeless shelters?
2. What is the evidence on transmission among PEH in different settings (e. g. in homeless shelters, at outreach events, when sleeping rough on the street)?
3. What are clinical and other health-related outcomes of the disease among PEH (e. g. measured by hospitalisation, ICU, ventilation, mortality)?
4. What is the evidence on the effects of lockdown measures and other non-pharmacological interventions on the health status of PEH?
5. What is the evidence on the effects of policies/strategies specifically enacted for PEH?

To cover unpublished research results, we searched pre-print repositories (medRxiv, bioRxiv), websites of 6 relevant institutions (see supplementary material 1) and the live map of COVID-19 evidence by the Norwegian Institute for Public Health.

A PICO framework was used to help define the inclusion/exclusion criteria: population was defined as PEH fitting the FEANTSA-ETHOS definitions; empirical quantitative, qualitative and mixed-methods studies, documenting exposure to SARS-Cov-2 or any policy measure or specific IPC strategies for PEH during the Pandemic were included; the general population was considered the standard for comparison, although a comparison group was not a criteria for inclusion; outcomes could be any health-related effect of SARS-Cov-2, including prevalence, incidence, evidence on transmission, development of disease, risk of severe course of disease, hospitalisation, ICU utilization, ventilation, mortality, etc. Effects of lockdown and other NPIs on the health status of PEH were also considered.

Empirical studies in English, indexed from December 1^st^ 2019 onwards, were included, and the searches were conducted on June 5^th^ and 9^th^ 2020. The table of contents of 4 relevant journals were also searched (Clinical Infectious Diseases, European Journal of Homelessness, The Lancet, Lancet Psychiatry).

### Data analysis

Titles and abstracts were screened by two independent researchers applying pre-defined inclusion/exclusion criteria. After screening 20%, the review team refined the criteria for the remaining articles. Full texts were obtained for all included studies and for those where no agreement could be established based on title/abstracts alone. For all included full text articles, backward and forward citation search was conducted (PRISMA Flowchart figure 1).

**Figure 1:**
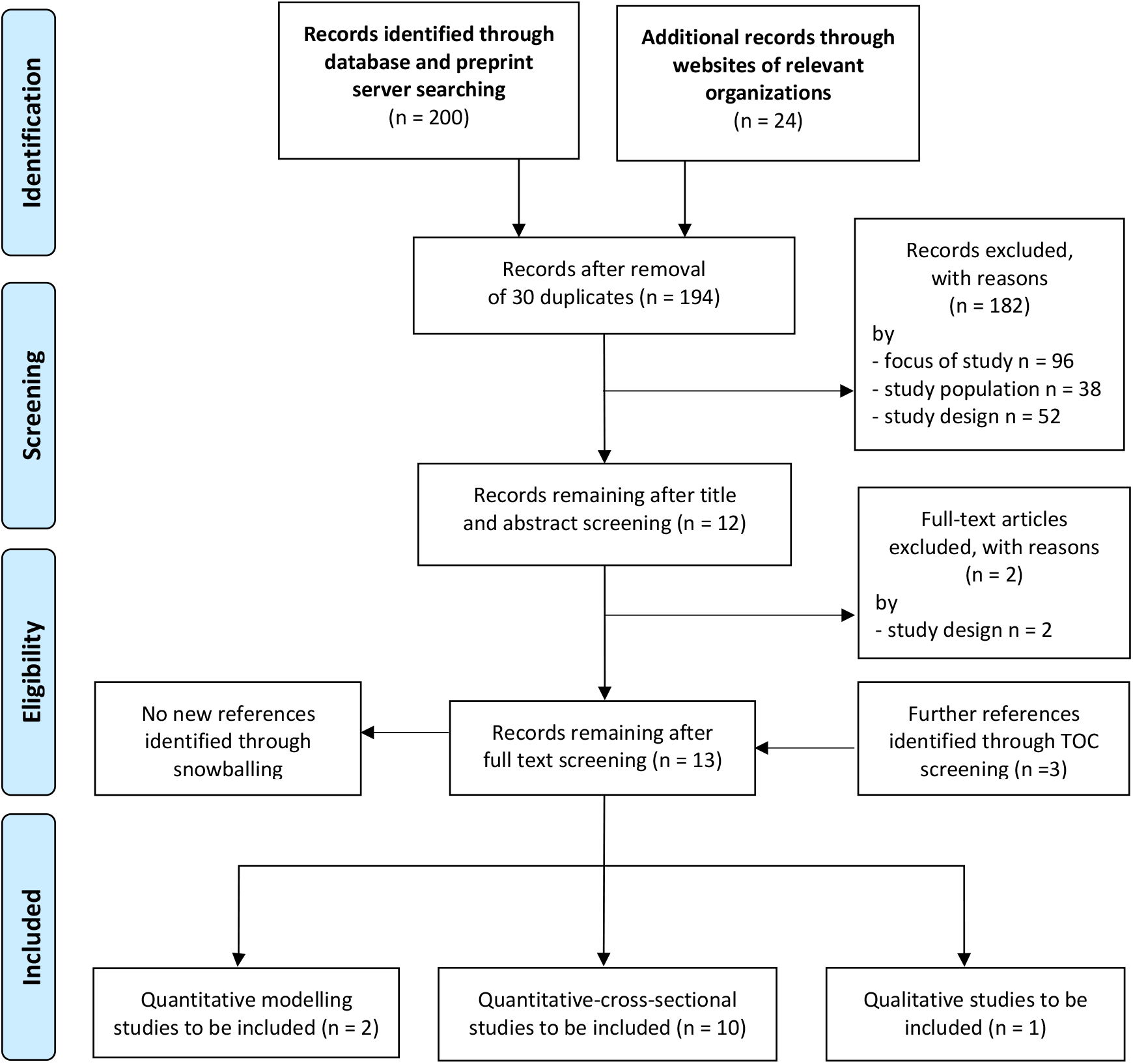
PRISMA 2009 Flow Diagrams.

We extracted relevant parameters from cross-sectional quantitative studies to synthesise a pooled measure of SARS-Cov-2 prevalence.

The extracted data was stratified based on the presence of an outbreak situation during data collection, either marked as such by authors or based on a prevalence of >=5%. Risk of bias and publication bias were assessed through funnel plots and regression-based statistical testing by Egger ^12,13^.

Stata V16^14^ with commands metaprop^15^ and meta^16^ were used in analysis.

## Results

After duplicates removal, 194 title and abstracts were eligible for screening. Twelve studies were included for full-text screening and ten eventually included in analysis. The screening of table of contents of specific journals, retrieved three additional studies.

Information was thus extracted from ten cross-sectional quantitative studies, two studies modelling various indicators and one qualitative study exploring experiences of women in unstable housing and previous criminal justice involvement during the pandemic (PRISMA Flowchart figure 1)^17^.

Eight of the cross-sectional quantitative studies were conducted in the USA^18–25^, one in Canada^26^ and one in France^27^. All studies collected data from March to early May 2020 during the first wave of the SARS-Cov-2 pandemic, except one^20^, conducted from April to August 2020.

All studies focused on people experiencing houselessness (i.e. with a place to sleep but temporary in institutions or shelter according to FEANTSA-ETHOS definitions^5,6^) with one study^21^ additionally testing people experiencing rooflessness. Nine articles reported PCR-based estimates of SARS-Cov-2 prevalence (with some offering additional data on other health or living conditions of homeless people), while one study^24^ researched shelter tenants’ knowledge and perception of COVID-19 and their adherence to behavioural rules. A total of six articles reported prevalence estimates for staff working at the homeless facilities^18,21–23,26,27^. Studies’ characteristics are presented in **Table 1**

**Table 1.** Summary of studies included.

| **Type of publication/**<br>**study** | **First author**<br>**(date of**<br>**publication)** | **ETHOS category of**<br>**homelessness** | **City (Country),**<br>**sample size PEH,**<br>**study period** | **Research objectives** | **Limitations as reported** |
| --- | --- | --- | --- | --- | --- |
| Peer-reviewed,<br>quantitative-cross-sectional | Bagget et al.<br>(Jun 2, 2020) | Not explicitly stated:<br>Houselessness | Boston (USA),<br>n = 408,<br>April 1-2, 2020 | SARS-CoV-2 testing of all residents from a single large homeless shelter in Boston, USA, exhibiting increasing numbers of COVID-19 cases | cross-sectional study at a single shelter in Boston where several symptomatic individuals had been removed through prior symptom screening or self-referrals to outside care |
| Peer-reviewed,<br>quantitative-cross-sectional | Imbert et al.<br>(Aug 2, 2020) | Not explicitly stated:<br>Houselessness | San Francisco (USA),<br>n = 255 (only 150 tested),<br>March 29-April 11, 2020 | To describe the lessons learned from the public health response to a COVID-19 outbreak that occurred | cross-sectional study at a single shelter in San Francisco; poor case interview completion rate and limited number of close contacts identified; at-risk population not completely represented; only 59% (n=155) of at-risk population (n=255) were tested; |
| Peer-reviewed,<br>quantitative-cross-sectional | Marquez et al.<br>(Oct 28, 2020) | Not explicitly stated:<br>Houselessness | San Diego (USA),<br>n = 2456,<br>April 16-August 5, 2020 | Preemptive testing was undertaken to identify potential asymptomatic residents, staff, or volunteers with the goal of preventing a potential community outbreak | Not enough info provided; |
| Peer-reviewed,<br>quantitative-cross-sectional | O'Shea et al.<br>(Jun 8, 2020) | Not explicitly stated:<br>Houselessness | Hamilton (Canada),<br>n = 104,<br>March 17-April 30, 2020 | To describe experience with shelter facility restructuring, daily symptom screening, and rapid testing to mitigate the risk of COVID-19 in the homeless shelter setting in Hamilton, Ontario, Canada | Potential selection bias: only those who were symptomatic through active screening within shelters. |
| Peer-reviewed,<br>quantitative-cross-sectional | Yoon et al.<br>(May 1, 2020) | Not explicitly stated:<br>Houselessness | Atlanta (USA),<br>n = 2326,<br>April 7-May 6, 2020 | To (1) determine the SARS-CoV-2 prevalence among clients living sheltered and | Cross-sectional of selected shelters in Atlanta; different specimen collection methods |

| Type of publication/<br>study | First author<br>(date of<br>publication) | ETHOS category of<br>homelessness | City (Country),<br>sample size PEH,<br>study period | Research objectives | Limitations as reported |
| --- | --- | --- | --- | --- | --- |
|  |  |  |  | unsheltered and homelessness service staff through viral testing;<br>(2) describe the clinical statuses of PEH and staff at the time of testing;<br>3) evaluate the sensitivity and specificity of symptom screening for COVID-19 detection;<br>(4) review shelter infection prevention and control (IPC) policies and provide recommendations to mitigate SARS-CoV-2 transmission | used; underestimation of people reporting symptoms. |
| Not peer-reviewed (MMWR*), quantitative-cross-sectional | Mosites et al. (Sep 8, 2020) | Not explicitly stated: Houselessness and rooflessness | Seattle, Boston, San Francisco, Atlanta (USA) n = 2326 (1690 sheltered and 636 unsheltered), March 27-April 15, 2020 | To test all shelter residents and staff members at each assessed facility, irrespective of symptoms | Cross-sectional study with some refusals acknowledged; Symptom information for persons tested was not consistently available and not included. |
| Not peer-reviewed (MMWR*), quantitative-cross-sectional | Tobolowsky, et al. (May 1, 2020) | Not explicitly stated: Houselessness | Seattle (USA), n = 299, March 30-April 8, 2020 | None explicitly stated. | Not all residents were present during the site visits. Thus, residents with SARS-CoV-2 infection could have been missed during the testing events or symptom screening. |
| Not peer-reviewed (pre-print), quantitative-cross-sectional | Henwood et al. (Apr 24, 2020) | PEH prioritized for Housing First programmes (permanent supportive housing) | Los Angeles (USA), n = 532, March 20-27, 2020 | To examine tenants' knowledge of COVID-19; perceived risk; pre-existing condition risk factors; consistency of handwashing and social distancing since the outbreak began; recent experiences of flu-like symptoms, and tenants' ability to shelter in place | Self-reported results. |
| Not peer-reviewed (pre-print), quantitative-cross-sectional | Ly et al.<br>(May 14, 2020) | Not explicitly stated: Houselessness | Marseille (France).<br>N = 411,<br>March 26-April 17, 2020 | To conduct a screening campaign among sheltered homeless individuals and compare them with asylum-seekers, other people living in precarious conditions and employees | Study population was not randomly and homogeneously recruited. |
| Not peer-reviewed (pre-print), quantitative-cross-sectional | Samuels et al.<br>(May 24, 2020) | Not explicitly stated: Houselessness | Providence (USA),<br>n = 299,<br>April 19-24, 2020 | To describe the varying prevalence of asymptomatic SARS-CoV-2 infection in congregate shelters and associated shelter characteristics and practices | At the time of study, many shelter residents who had tested positive were already housed in a hotel, which likely led to an underestimate of true prevalence in the unhoused population; Testing done at the shelters with transient residents only reflects those staying there on the night of testing, and not intermittent users. |
| Grey-literature report, modelling | Culhane et al.<br>(Mar 27, 2020) | US Department of Housing and Urban Development definition: "living in a homeless shelter or a place not fit for human habitation" | USA, modelling for all homeless (n=575.000), Estimated during March, 2020 | To establish the potential mortality and hospitalization costs of inaction along with estimating the funding needs associated with a comprehensive plan of action | Application of homogenous rates (infections, hospitalizations, mortality) as assumptions for the whole country (US). Modelling based on a single point-in-time estimate of homelessness. |
| Peer-reviewed, modelling | Lewer et al.<br>(Sep 23, 2020) | Core homelessness groups in England, which includes people sleeping rough, living in tents or cars, squatting, living in temporary homeless hostels, and sofa-surfing | England, modelling for all homeless, February-March, 2020 | To estimate the avoided deaths and health-care use among people experiencing homelessness during the so-called first wave of COVID-19 in England and the | Uncertainty about the size and structure of homeless population; assumed no mixing between subgroups of homeless; did not vary the degree of infectiousness or duration of disease states by |
|  |  |  |  | potential impact of COVID-19 on this population in the future | severity in models; treated homeless population as static; |
| Peer-reviewed, qualitative | Ramaswamy et al.<br>(May 8, 2020) | Not explicitly stated:<br>Houselessness | USA, 35 participants from a cohort of criminal justice-involved women's health and cervical cancer risk (total cohort size is 474), March-April, 2020 | To describe the experiences of women living in the community who simultaneously negotiate criminal justice involvement and COVID-19 in three urban areas | Qualitative assessment |

\*MMWR = Morbidity and Mortality Weekly Report by CDC

### Prevalence of SARS-Cov2 in homeless shelters

We found 9 quantitative articles reporting PCR-based SARS-Cov-2 prevalence estimates in PEH and 6 studies reporting prevalence estimates for shelter staff members. To adjust for the varying settings and contexts, we pooled the measures in a random-effects meta-analysis model for PEH (n=7107) and staff (n=1375). To consider the difference in nature of investigating an outbreak^18,19,22,23,25,27^ and testing for baseline estimation^18,20,21,26,27^ in our meta-analysis, we sub-pooled the measures accordingly.

For PEH, the pooled SARS-Cov-2 prevalence estimate was 29·49% (95% Confidence Interval, 95%CI=16·44%-42·55%, I^2^=97·79%, p=0·00) in the context of outbreak situations. As a sensitivity analysis, we removed an outlier that could reflect a super-spreading event (Imbert et al.,^22^ 67·33%, 95%CI=59·48%-74·32%), resulting in a pooled prevalence of 23·32% (95%CI=12·12%-34·53%, I^2^=96·79%, p=0·00). For baseline estimation, the pooled prevalence was 2·14% (95%CI=1·02%-3·27%, I^2^=83·80%, p=0·00) (figure 2).

**Figure 2:**
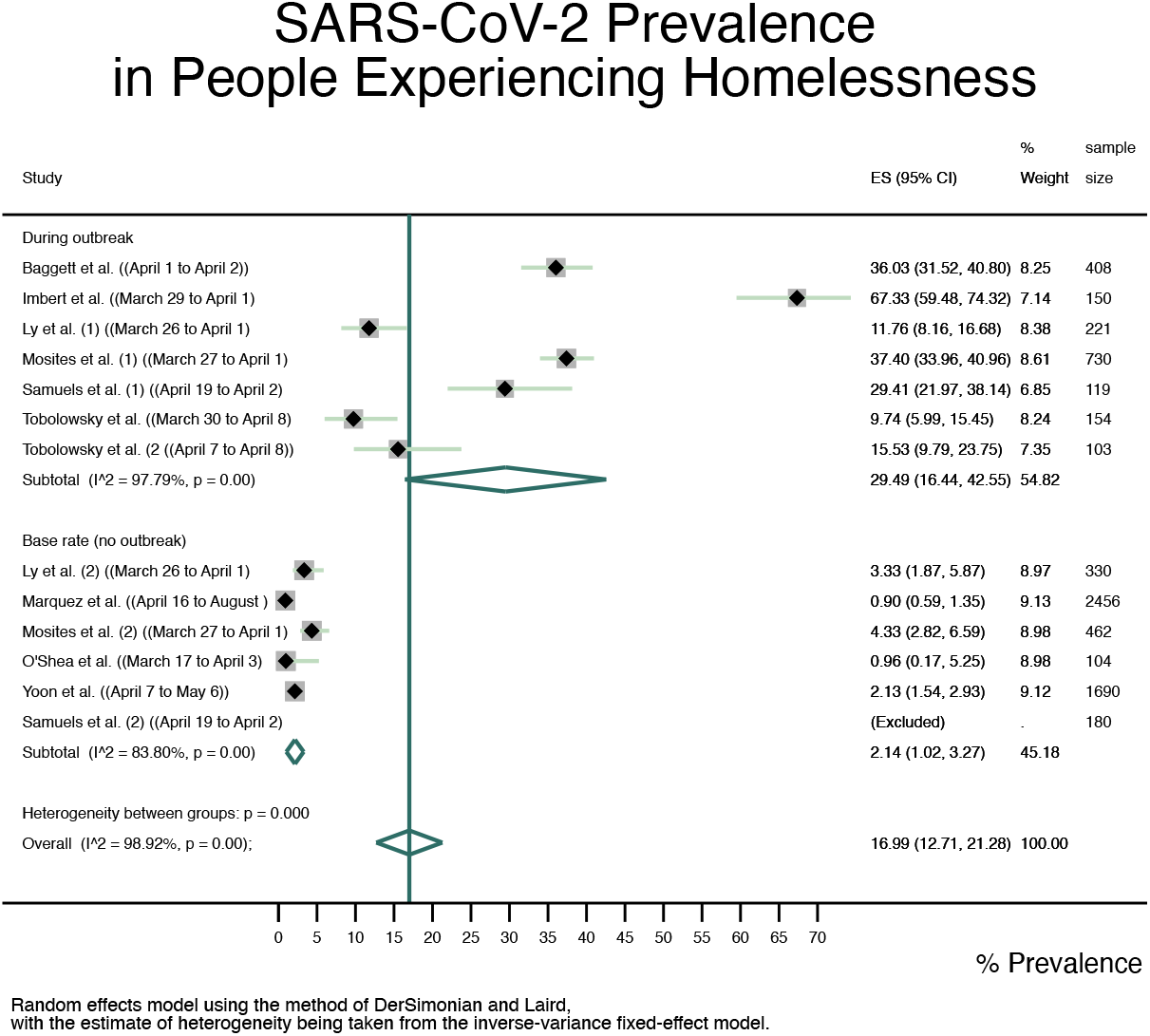
Forest plot of SARS-Cov-2 prevalence pooled by outbreak situation and in total for PEH.

For staff members, the random-effect meta-analysis resulted in a pooled prevalence of 15·18% (95%CI=8·95%-21·42%, I^2^=65·76%, p=0·02) during an outbreak situation and 1·72% (95%CI=0·31%-3·12%)) for baseline estimation (figure 3).

**Figure 3:**
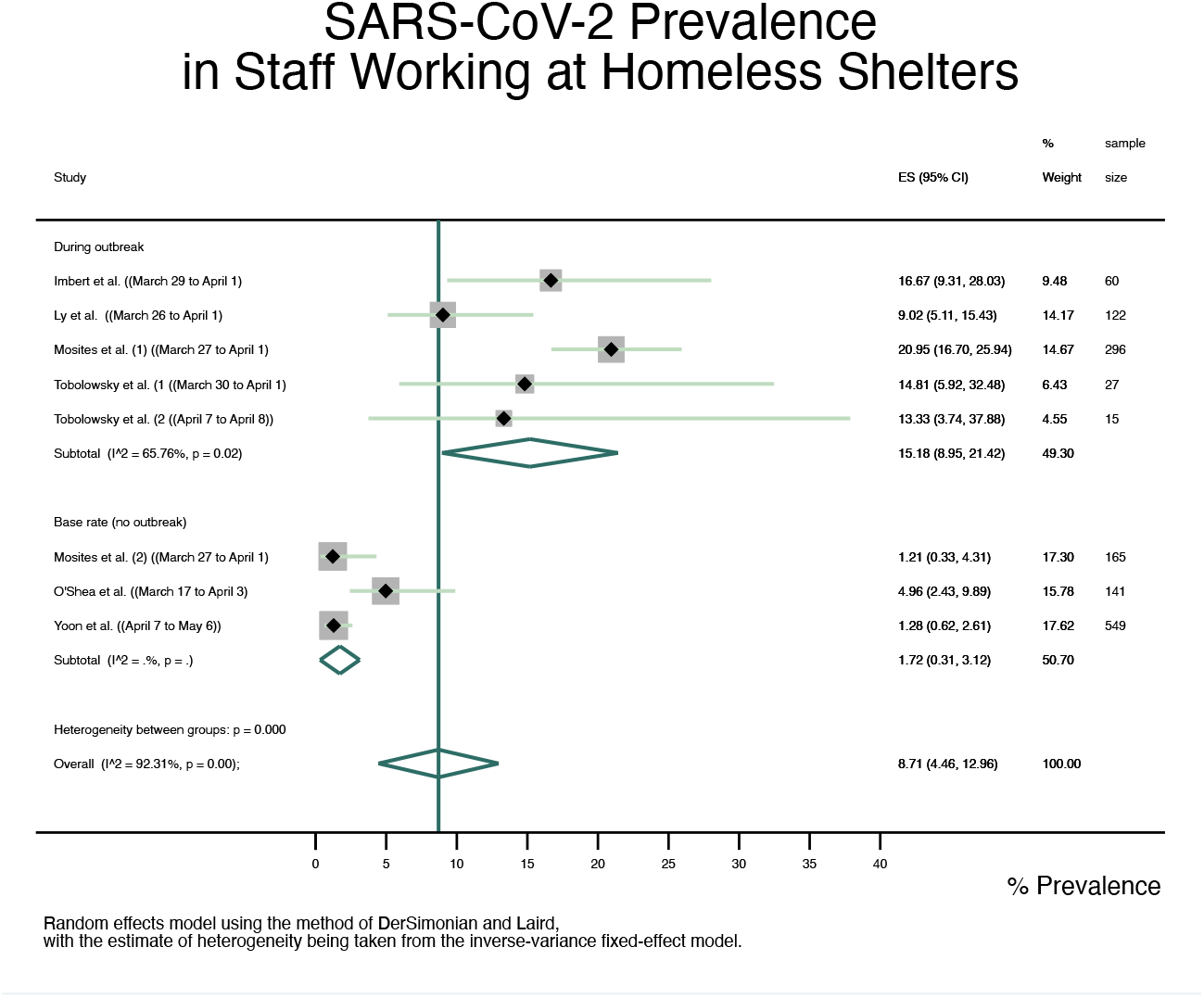
Forest plot of SARS-Cov-2 prevalence pooled by outbreak situation and in total for staff.

### Transmission among PEH

While all studies tested PEH in homeless shelters, one study by Yoon et al.^21^ conducted additional testing of clients living unsheltered during homeless outreach service events (e.g. meal services). This resulted in a prevalence of 0·5% (3 out of 636) in the unsheltered PEH.

### Clinical and health-related outcomes among PEH

Five articles^19,21,24,25,27^ presented data on clinical outcomes of PEH, ranging from respiratory symptoms to previous mental health diagnosis and other somatic issues (e.g. chronic kidney or liver disease), and one article^22^ provided data on emergency department visits, hospitalization and mortality. Four articles^19,21,22,25^ provided data specifically for PEH testing positive: specific symptoms (cough, shortness of breath, fever) were either uncommon or did not differ significantly from the overall tested population of PEH. Neither did comorbidities^25^. Imbert et al.^22^ used a city-wide administrative database and reported 12% (n=12) of those tested positive to have had a treatment and release emergency department visit, 8% (n=8) a hospitalization and 1% (n=1) died.

Comparing to RT-PCR, Yoon et al. calculated sensitivity and specificity of symptom screening, stating that fever, cough, or shortness of breath was 14% sensitive and 89% specific for identifying COVID-19 cases when reported in the previous day, changing to 24% and 85% when reported in the previous week. An expansion to any symptom reported resulted in no improvement of sensitivity, but a loss in specificity (last week: 81%; last day: 87%).

Two studies modelled different health-related outcomes among PEH with one modelling conducted for England^28^ and one for the USA^29^.

Lewer et al. (2020)^28^ aimed to estimate the impact of COVID-19 on the population of PEH of England and estimate the reduction in mortality and healthcare use that could result from a targeted intervention compared with a “do nothing” scenario.The authors used a Markov chain model with a modified susceptible-exposed-infectious-recovered (SEIR) model, assuming a basic reproductive rate of 1·4 in the baseline scenario, a latency, incubation and infectiousness period of 2·5 days, 5 days, and 6·5 days respectively, and infection fatality ratio for patients of 4.8% for those homeless defined as vulnerable (37%) and 0·6% for the remaining population. For the latter scenario, the authors estimated that 15448 cases of COVID-19 could occur among the 35817 homeless and 10748 people sleeping rough in England, resulting in a cumulative incidence of 34%.

This incidence could be reduced to 21% (i.e. 9934 cases) with a targeted intervention by the housing and health authorities in England. The authors also estimated a total of 364 death if nothing was implemented for PEH in England but a reduction to 200 deaths with the intervention.

The report by Culhane et al (2020)^29^ aimed to estimate the mortality, hospitalisation costs and system capacity to manage the impact of COVID-19 pandemic on the existing population of PEH in the USA. Based on data on the age distribution of PEH from two large cities (Los Angeles and New York) and on studies that older homeless populations bear health risks comparable to those of individuals 15-20 years old, the authors estimated that there were 4·3% PEH across the USA (corresponding to 21295 people, ranging from 2·4% to 10·3%) that could require hospitalization assuming an infection rate reaching 40%. They also estimated that critical care needs could range from 0·6% to 4·2% for the homeless and fatality rates could range from 0·3% to 1·9% (central estimate 0·7% or 3454 deaths).

### Non-pharmaceutical interventions (NPI)

We could not identify articles quantifying the effects of NPIs on PEH. However, one qualitative study^30^ conducted unstructured interviews through telephone or social media conversations with 35 participants from a US cohort study of criminal justice-involved women to explore how they were faring during this period. Thirteen of the interviewees reported some form of homelessness, and for some of the women their unstable living situations prevented them from being able to follow the social distancing recommended.

### Infection protection and control (IPC) strategies in homeless shelters

We identified no study quantifying the effectiveness of strategies for IPC among PEH. Five papers^23,25,26,28,29^ discussed strategies for IPC more generally, and one study^21^ actively investigated these at 9 shelters. Yoon et al.^21^ describe different pandemic preparation measures, e.g., 55·6% of shelters (5 in 9) discontinued taking in new clients, 77·8% (7 in 9) created separate isolation areas for PEH with suspected infection and 55·6% (5 in 9) increased the spacing between individual beds.

Across all studies, the major strategies discussed as important and necessary were comprehensive and rapid testing, expansion of non-congregate supportive housing for PEH and, specific strategies for IPC to be taken in congregate settings, e.g. increased spacing between beds and/or limited staff rotation. O’Shea et al. further discussed potential benefits in collaborating with regional laboratories for quick turnaround of test results which enable the isolation of the infected.

Furthermore, Henwood et al^24^ reported that 39% (207 of 532) of the tenants of supportive housing in Los Angeles considered themselves to be at serious risk for COVID-19 due to a pre-existing condition, 59% (316 of 532) had a registered mental health diagnosis and 55·3% (294 of 532) believed they can shelter in place if needed.

Funnel plots of studies reporting SARS-Cov-2 prevalence and corresponding standard errors are presented in figure 4. The Egger test suggests absence of significant publication bias in all strata except for the overall prevalence of PEH, which disappears after stratification by outbreak situation (supplementary material 3), supporting differentiation between baseline estimation and outbreak investigation.

Risk of bias assessment is presented in table 2.

**Table 2:** Risk of bias assessment of included studies.

| Article | Publication status | Clear research question | Generalizable to all PEH | Generalizable to study setting | Correct classification of exposure | Correct classification of outcome | Controlling for confounders | Transparency and justification of structural Assumptions | Transparency and justification of input parameters | Sufficient technical documentation |
| --- | --- | --- | --- | --- | --- | --- | --- | --- | --- | --- |
| Bagget | peer-reviewed | + | - | + | N/A | + | N/A | N/A | N/A | N/A |
| Imbert | peer-reviewed | + | - | + | N/A | + | N/A | N/A | N/A | N/A |
| Marquez | peer-reviewed | + | - | + | N/A | + | N/A | N/A | N/A | N/A |
| O'Shea | peer-reviewed | + | - | + | N/A | + | N/A | N/A | N/A | N/A |
| Yoon | peer-reviewed | + | - | + | N/A | + | N/A | N/A | N/A | N/A |
| Mosites | Not peer-reviewed (MMWR) | + | - | ? | N/A | + | N/A | N/A | N/A | N/A |
| Tobolowsky | not peer-reviewed (MMWR) | 0 | - | + | N/A | + | N/A | N/A | N/A | N/A |
| Henwood | pre-print | + | - | + | + | + | + | N/A | N/A | N/A |
| Ly | pre-print | + | - | + | N/A | + | N/A | N/A | N/A | N/A |
| Samuels | pre-print | + | - | ? | N/A | + | N/A | N/A | N/A | N/A |
| Ramaswamy | Peer-reviewed | + | - | + | N/A | N/A | N/A | N/A | N/A | N/A |
| Culhane | grey-literature | + | - | N/A | N/A | N/A | N/A | - | 0 | 0 |
| Lewer | Peer-reviewed | + | - | N/A | N/A | N/A | N/A | + | + | + |

“+” good
“0” medium
“-” bad
“?” – unknown
“N/A” not applicable
MMWR = Morbidity and Mortality Weekly Report by CDC

## Discussion

In this systematic review including a total of 13 empirical studies, we identified a baseline SARS-Cov-2 prevalence of 2·14% (95% Confidence-Interval, 95%CI=1·02-3·27%) among PEH in homeless shelters, and 1·72% (95%CI=0·31-3·12%) among staff. In case of an outbreak situation, these estimates increase to 29·49% (95%CI=16·44-29·55%) among PEH and 15·18% (95%CI= 8·95-21·42%) among staff. Among unsheltered PEH, one study reported a prevalence of 0·5%.

Regarding health-related outcomes, specific symptoms, e.g. cough, shortness of breath or fever, were either uncommon or did not differ significantly from the overall tested population of PEH^19,21,22,24,25,27^, neither did comorbidities^25^. One study reported 12% treat-and-release emergency department visits, 8% hospitalization and 1% death among those who tested positive, based on a city-wide administrative database^22^ without specifying the time of follow-up. Further, one study determined reporting of fever, cough or shortness of breath to have been 14-24% sensitive and 85-89% specific compared to RT-PCR testing^21^.

Based on one qualitative study, unstable living situations were reported to be preventing criminal-justice involved women from following social distancing practices recommended.

Regarding IPC strategies, 6 studies^21,23,25,26,28,29^ discussed comprehensive and rapid testing including collaboration with regional laboratories for quick turnaround of positive tests, expansion of non-congregate housing and individual strategies in the shelters, e.g. spacing between beds and limited staff rotation, as critical measures

The modelling studies reported potential SARS-Cov-2 infection among PEH of 21295 (4% of the general population) in the USA^29^ and 22933 (cumulative incidence of 49.3% among PEH) in England^28^ during the first wave if no intervention measures had been taken. Even if incidence of SARS-Cov-2 remains low in the general population, Lewer et al^28^ conclude that an outbreak among PEH can lead up to 11566 infections if no prevention measures are in place.

Although excluded from this review, we found several reports retrieved from the relevant websites searched providing IPC strategies specifically for staff working in homeless shelters (supplementary material 4). These strategies should be adapted to the respective context and enacted to mitigate the spread of SARS-Cov-2 infection among this vulnerable population group. We further identified important research gaps to be addressed by future research (Panel 1)

### Quality of Evidence

Studies providing prevalence estimates were based on PCR testing and some included testing of staff for a complete picture of SARS-Cov-2 infection in shelters. Overall, the quality of evidence was low. Most articles provide little detail on how and why shelters were sampled. Studies may have missed infected patients who do not have enough viral particles yet to be detected in PCR. Only two articles conducted follow-up testing^21,23^. The recruitment of participants was based on accommodation in homeless shelters with no systematic surveying of housing situation and residence type in shelter. This may be of special importance considering the diverse background of PEH and their everyday life, because not all shelters offer accommodation during the day. Finally, all identified studies were based in high-income countries raising questions of generalizability especially to middle- and lower-income countries that differ in population demographics, cultural and social dimensions. Further details about the risk of bias are provided as supplementary material 3.

### Strengths and limitations

Besides major databases, we searched pre-print repositories and websites of relevant institutions for unpublished manuscripts and consulted the Covid-19 evidence live map of the Norwegian Institute of Public Health. Further, we have used forward and backward reference screening on all identified articles and screened the TOCs of journals deemed relevant for potential publications during the screening, extraction, and analysis phases. All steps of the review have been conducted by two reviewers for quality assurance. Finally, we tested for publication bias, both visually through funnel plotting as well as statistically based on Egger^12,13^. Additionally, danger of publication bias might be low as we are investigating prevalence (compared to measures of effect) during the onset of a pandemic of global relevance.

We have used English search terms only, thus might have missed relevant studies in other languages. This could explain why only one study from a non-English language country was identified. Our findings may not be generalizable to other settings than the ones where the studies were conducted, as droplet or potentially airborne transmission may be different in other contexts of social and cultural communication and interaction. Third, even though we searched several databases and repositories, we might not have detected all relevant studies considering the amount of evidence being generated during the current pandemic. Additionally, we cannot rule out the possibility of publication bias, particularly regarding the baseline prevalence among staff since estimates for staff were not part of these studies’ initial objectives.

## Conclusions

As the quality of included studies varies, findings should be considered with caution. Yet, they clearly suggest that PEH are at high risk of being infected with SARS-Cov-2, especially in case of outbreaks in shelters. Potential pathways for transmissions deserving further investigation are the “mobile nature of the community and use of multiple homeless service sites among residents”^23^, crowded sleeping arrangements and difficulties in adhering to recommended behavioural interventions due to lack of resources and private space. Furthermore, PEH face serious mental health challenges^24^ and economic difficulties that prevent them from accessing preventive material and maintaining physical distancing in general^30^.

The nature and direction of the infectious disease spread is unclear but staff working at shelters are at increased risk as well, reinforcing the importance of adherence to strategies of IPC in congregate shelters and the expansion of provision of non-congregate housing for PEH.

Due to the absence or low prevalence of symptoms (e.g. fever) at the time of diagnosis of SARS-Cov-2 among PEH^19,21,25,27^, symptom screening alone is not suitable, i.e. not sensitive enough for adequately capturing the extent of disease transmission in such high-risk settings. Instead, comprehensive and early testing for timely identification and isolation of the infected, combined with collaboration with regional laboratories for quick turnaround of positive test results might be important strategies to break the virus transmission^21,26^.

### Panel 3 Research gaps identified

**(A) Lack of data for evaluation of NPIs effectiveness and (un)intended health consequences**

(1) lack of reliable and separate data on PEH within Covid-19 notification systems

(2) lack of a comprehensive overview of NPIs and shelter strategies targeting PEH

(3) lack of understanding of infectious disease spread among PEH, e.g. in homeless shelters

**(B) Targeted research into people experiencing rooflessness**

While PEH may be deemed as a hard-to-reach population in general, it is essential to differentiate those living in shelters from those sleeping rough. People experiencing rooflessness may be at a less risk of SARS-Cov-2 infection as long as they do not seek congregate shelter but at the same time, they may be the ones experiencing more heavily the challenges of social distancing to mental and social wellbeing associated with the imposed lockdowns.

**(C) Lack of qualitative research into living realities of PEH**

More qualitative research is needed to better understand living realities and particular needs of PEH during the pandemic. This can further kick-start the participation of PEH in the research process and increase

**(D) Timely updates of the review**

With the current infection dynamics and speed of research, timely updates of synthesised information is essential.

## Data Availability

Detailed review lists can be provided by the corresponding author upon reasonable request.

## Acknowledgements

None.

## Funding

This research did not receive any specific grant from funding agencies in the public, commercial, or not-for-profit sectors.

Data availability statement

Detailed review lists can be provided by the corresponding author upon reasonable request.

## Authors’ contributions

Conceptualisation: AM, DC, KB

Data curation / Search: Embase - SR; WHO Covid19 - DC; WoS - SR; pre-print repositories - DC, AM; websites - DC; NIPH live map - AM; Snowballing: JS

Title and abstract screening: AM, DC Full-text screening: AM, DC

Quality appraisal: AM, DC, JS, SR Data extraction: AM, DC

Data-synthesis: AM, DC

Writing of first and final draft: AM, DC

Revision for important intellectual content: KB, JS, SR

## Ethics statement

The review is based on published literature, no ethical clearance was required.

## Supplementary Material

### 1 Search strategy

1 – Search strategy
| Search syntax development (EMBASE) on 5 June 2020 |  |  |
| --- | --- | --- |
| Search-id | Search terms | No of hits |
| #11 | #4 AND #9 AND #10 | 45 |
| #10 | [12-1-2019]/sd | 3,069,260 |
| #9 | #5 OR #6 OR #7 OR #8 | 40,949 |
| #8 | (homeless:ab,ti,kw,de OR shelterless:ab,ti,kw,de OR unsheltered:ab,ti,kw,de OR rooflessness:ab,ti,kw,de OR houselessness:ab,ti,kw,de OR campsite:ab,ti,kw,de OR destitute:ab,ti,kw,de OR dispossess*:ab,ti,kw,de OR outcast:ab,ti,kw,de OR 'social outcast':ab,ti,kw,de OR vagabond:ab,ti,kw,de OR vagrant:ab,ti,kw,de OR derelict:ab,ti,kw,de OR itinerant:ab,ti,kw,de OR 'down-and-out':ab,ti,kw,de OR 'down and out':ab,ti,kw,de OR 'rough sleeping':ab,ti,kw,de OR ((homeless:ab,ti,kw,de OR shelterless:ab,ti,kw,de OR unsheltered:ab,ti,kw,de OR destitute:ab,ti,kw,de OR dispossess*:ab,ti,kw,de) AND (people:ab,ti,kw,de OR person:ab,ti,kw,de OR women:ab,ti,kw,de OR men:ab,ti,kw,de OR children:ab,ti,kw,de OR families:ab,ti,kw,de OR family:ab,ti,kw,de)) OR ((inadequate*:ab,ti,kw,de OR insecure*:ab,ti,kw,de OR precarious*:ab,ti,kw,de OR unstabl*:ab,ti,kw,de) AND housing:ab,ti,kw,de)) AND ([embase]/lim OR [medline]/lim OR [pubmed-not-medline]/lim) | 13,784 |
| #7 | 'housing'/exp AND ([embase]/lim OR [medline]/lim OR [pubmed-not-medline]/lim) | 25,630 |
| #6 | 'homeless person'/exp AND ([embase]/lim OR [medline]/lim OR [pubmed-not-medline]/lim) | 1,987 |
| #5 | 'homelessness'/exp AND ([embase]/lim OR [medline]/lim OR [pubmed-not-medline]/lim) | 10,979 |
| #4 | #1 OR #2 OR #3 | 41,649 |
| #3 | ((('corona virus':ab,ti,kw,de OR 'corona viruses':ab,ti,kw,de OR coronavir*:ab,ti,kw,de OR coronavirus*:ab,ti,kw,de OR betacoronavirus*:ab,ti,kw,de) AND (novel:ab,ti,kw,de OR 2019:ab,ti,kw,de OR wuhan:ab,ti,kw,de OR huanan:ab,ti,kw,de OR hubei:ab,ti,kw,de) OR 'new coronavirus':ab,ti,kw,de OR 'covid-19':ab,ti,kw,de OR covid19:ab,ti,kw,de OR 'sars coronavirus 2':ab,ti,kw,de OR 'severe acute respiratory syndrome coronavirus 2':ab,ti,kw,de OR ncov:ab,ti,kw,de OR 2019ncov:ab,ti,kw,de OR ncov2019:ab,ti,kw,de OR 'sars-cov-2':ab,ti,kw,de OR 'sars-cov2':ab,ti,kw,de OR sarscov19:ab,ti,kw,de OR 'sars cov19':ab,ti,kw,de OR 'sars cov 19':ab,ti,kw,de OR 'hcov 19':ab,ti,kw,de OR 'wn cov':ab,ti,kw,de) AND ([embase]/lim OR [medline]/lim OR [pubmed-not-medline]/lim) | 19,482 |
| #2 | 'coronavirus infection'/exp AND ([embase]/lim OR [medline]/lim OR [pubmed-not-medline]/lim) | 14,313 |
| #1 | 'coronaviridae'/exp AND ([embase]/lim OR [medline]/lim OR [pubmed-not-medline]/lim) | 21,926 |

| Search syntax development (WoS CC) on 5 June 2020 |  |  |
| --- | --- | --- |
| Indexes=SCI-EXPANDED, SSCI, A&HCI, ESCI Timespan=All years |  |  |
| Search-id | Search terms | No of hits |
| #4 | #1 AND #2 Timespan=2019-2020 | 11 |
| #3 | #1 AND #2 | 14 |
| #2 | TS = (homeless OR shelterless OR unsheltered OR rooflessness OR houselessness OR campsite OR destitute OR dispossess* OR outcast OR "social outcast" OR vagabond OR vagrant OR derelict OR itinerant OR "down-and-out" OR "down and out" OR "rough sleeping" OR ((homeless OR shelterless OR unsheltered OR destitute OR dispossess*) AND (people OR person OR women OR men OR children OR families OR family)) OR ((inadequate* OR insecure* OR precarious* OR unstabl*) AND housing)) | 33,134 |
| #1 | TS = (("corona virus" OR "corona viruses" OR coronavir* OR coronavirus* OR betacoronavirus*) AND (novel OR 2019 OR Wuhan OR Huanan OR Hubei) OR "new coronavirus" OR "COVID-19" OR COVID19 OR "SARS coronavirus 2" OR "severe acute respiratory syndrome coronavirus 2" OR nCoV OR 2019nCoV OR nCoV2019 OR "SARS-CoV-2" OR "SARS-CoV2" OR SARSCoV19 OR SARS-CoV19 OR SARS-CoV-19 OR HCoV-19 OR WN-CoV OR "corona vir*" OR "coronavir*" OR "betacoronavir*" OR "severe acute respiratory syndrome coronavirus") | 21,307 |

| Others |  |  |  |
| --- | --- | --- | --- |
| Source | Search expression | Hits | Day |
| WHO COVID research:<br>https://search.bvsalud.org/global-literature-on-novel-coronavirus-2019-ncov/advanced/?lang=en | (tw:(homeless)) OR<br>(tw:(houseless)) OR<br>(tw:(roofless)) | 24 | 05 June 2020 |
| www.medrxiv.org | Homeless OR houseless<br>OR roofless | 69 | 5 June 2020 |
| www.biorxiv.org | Homeless OR houseless<br>OR roofless | 51 | 5 June 2020 |
| NIPH Live map COVID-19 evidence:<br>https://www.nornesk.no/forskningskart/NIPH_mainMap.html | Check map | 3 | 5 June 2020 |
| FEANTSA:<br>https://www.feantsa.org/en |  | 0 | 9 June 2020 |
| EUPHA:<br>https://eupha.org/ |  | 0 | 9 June 2020 |
| European Observatory on Homelessness:<br>https://www.feantsaresearch.org/ | Includes repository from the *European Journal of Homelessness* and *Comparative Studies on Homelessness* | 0 | 9 June 2020 |
| Pathway/<br>Faculty of Homeless and Inclusion Health:<br>https://www.pathway.org.uk/ |  | 2 | 9 June 2020 |
| Homeless Link:<br>https://www.homeless.org.uk/ |  | 14 | 9 June 2020 |
| Homeless Hub Canada:<br>https://www.homelesshub.ca/ | Publications (Reports) | 5 | 9 June 2020 |

| Snowballing |  |  |
| --- | --- | --- |
| Reference screening (backward) | Hits | Eligible/New |
| Baggett JAMA 2020 | 2 | 0/0 |
| Henwood preprint 2020 | 14 | 2/0 |
| Lewer preprint 2020 | 39 | 2/0 |
| Ly preprint 2020 | 15 | 3/0 |
| Mosites MMWR 2020 | 5 | 0/0 |
| Ramaswamy HE&B 2020 | 11 | 0/0 |
| Samuels preprint 2020 | 5 | 2/0 |
| Tobolowski | 9 | 0/0 |

| Citation screening (forward) |  |  |
| --- | --- | --- |
| Baggett JAMA 2020 | 7 | 0/0 |
| Henwood preprint 2020 | - | - |
| Lewer preprint 2020 | - | - |
| Ly preprint 2020 | - | - |
| Mosites MMWR 2020 | 1 | 0/0 |
| Ramaswamy HE&B 2020 | - | - |
| Samuels preprint 2020 | - | - |
| Tobolowski | 1 | 0/0 |

| PubMed search syntax for daily result updates |
| --- |
| Search: (((("covid-19"[nm] OR "severe acute respiratory syndrome coronavirus 2"[nm]) OR ((Coronavirus[mh] OR "Coronavirus Infections"[mh] OR Coronaviridae[mh:noexp] OR "Coronaviridae Infections"[mh:noexp] OR "corona virus"[tw] OR "corona viruses"[tw] OR coronavir*[tw] OR coronavirus*[tw] OR betacoronavirus*[tw]) AND (novel[tw] OR 2019[tw] OR Wuhan[tw] OR Huanan[tw] OR Hubei[tw] OR "new coronavirus"[tw] OR "COVID19"[tw] OR "COVID19"[tw] OR "SARS coronavirus 2"[tw] OR "severe acute respiratory syndrome coronavirus 2"[tw] OR nCoV[tw] OR 2019nCoV[tw] OR nCoV2019[tw] OR "SARSCoV-2"[tw] OR "SARS-CoV2"[tw] OR SARSCoV19[tw] OR "SARS-CoV19"[tw] OR "SARS-CoV19"[tw] OR "HCoV-19"[tw] OR "WN-CoV"[tw]))) AND (("2019/12/01"[Date - Entry] : "3000"[Date - Entry])) AND (("homeless persons"[mh] OR homeless*[tw] OR houseless*[tw] OR roofless*[tw]) OR ((inadequate*[tw] OR insecure*[tw] OR precarious*[tw] OR unstabl*[tw]) AND (housed[tw] OR housing[tw])))) |

### 2 Discussing potential publication bias

**Figure SM2:**
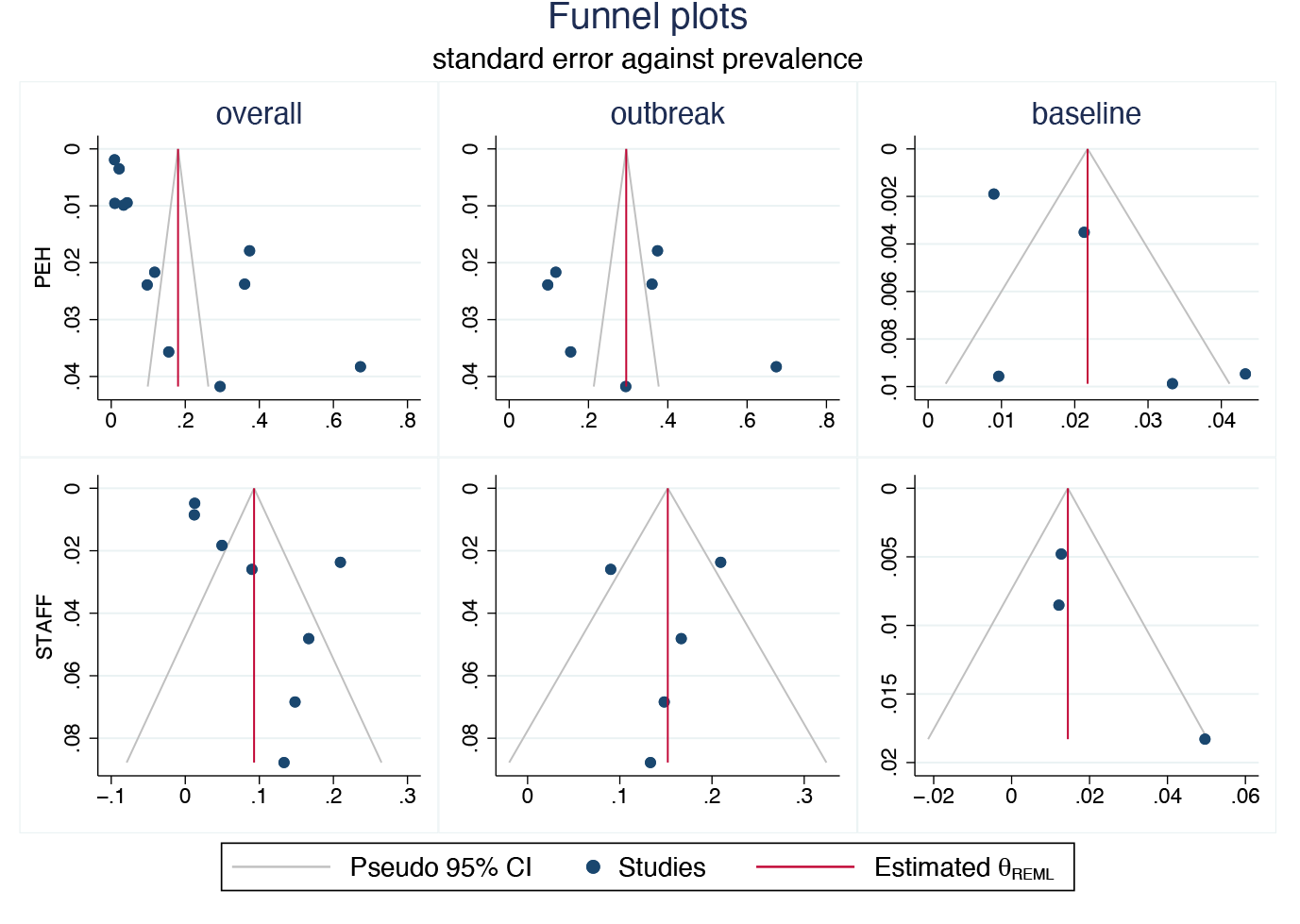
Forest plots of standard errors against prevalence, stratified outbreak situation for PEH and for staff.

**Table SM2:** Regression-based Egger test for publication bias effects Random-effects model H0: β1 = 0; no small-study effects.

**H0: $\beta_1 = 0$ ; no small-study effects**
| | $\beta_1$ | SE |
| --- | --- | --- |
| **PEH** |  |  |
| overall | **10.65\*\*** | 3.362 |
| base rate | 2.15 | 1.560 |
| outbreak | 6.95 | 9.097 |
| **Staff** |  |  |
| overall | 1.93 | 1.076 |
| base rate | 2.23 | 1.324 |
| outbreak | -0.22 | 1.461 |

95% confidence intervals in brackets;
\* $p < 0.05$ ;
\*\* $p < 0.01$ ;
\*\*\* $p < 0.001$ ;

### 3 Risk of bias assessment

We conducted a separate risk of bias assessment for all included articles. Summarizing the quantitative cross-sectional studies, all articles had a clear population by study design as a majority were conducted in homeless shelters, stated clear research objectives and, if reporting on prevalence, used PCR testing. Yet, few studies explicitly stated if data collection was conducted during a suspected outbreak situation or with the aim to provide baseline estimates and only 2 studies engaged in follow-up testing, one after 1 week^1^ and one after 3-4 weeks^2^ to investigate the full extent of a potential outbreak. As expected from studies conducted at an early stage of the pandemic, the majority of results are descriptive in nature.

The qualitative article^3^ offered a first insight into conversations with criminal justice-involved women yet offered little details on study design (including questions on how conversations were guided), details on interviewers and implications for the way interviews progress as well as ethical concerns. Further, the article does not provide information to check quality criteria in qualitative research (e. g. intersubjectivity), nor explores the interview findings in detail.

The two modelling studies offered insights into their assumptions and results, although the included grey-literature report^4^ lacks several details about assumptions taken. It further uses data from registers from two cities (Los Angeles and New York) to extrapolate the age range of homeless groups (aged 25+) for all US regions. The overall size of the homeless population is based on a “point-in-time” estimate, and the assumption of a 40% infection rate is based on existent literature (not necessarily from the US). The authors state using “nonlinear regression techniques” for their estimation, although a concrete model is not specified.

The modelling approach taken by Lewer et al.,^5^ describes all assumptions taken and provides concrete information of studies and conditions where initial assumptions were extrapolated from (e. g. using demographic profile of a homeless charity in London, based on census of providers of accommodations conducted during 2019, an official count of people sleeping rough, data from a multi-agency database for people sleeping rough, and data from night shelters in London). Furthermore, all the details from the actual modelling conducted (with respective syntax) are provided as additional material accessible with the publication in open access repository.

## 4 List of reports found with strategies for infection protection and control and relevant information about homelessness and Covid-19

Amanda Buchnea, Mary-Jane McKitterick, David French (2020). Summary Report: Youth Homelessness and COVID-19: How the youth serving sector is coping with the crisis. Toronto, ON: Canadian Observatory on Homelessness Press and A Way Home Canada. Availabe from: https://www.homelesshub.ca/coh-awh-youth-survey-covid19-2020

Leilani Farha, Kaitlin Schwan (2020). A National Protocol for Homeless Encampments in Canada: A Human Rights Approach. UN Special Rapporteur on the Right to Housing. Available from: https://www.homelesshub.ca/resource/human-rights-approach-national-protocol-homeless-encampments-canada%C2%A0

Keith Ahamad, Paxton Bach, Rupi Brar, Nancy Chow, Neasa Coll, Miranda Compton, Patty Daly, Nadia Fairbairn, Guy Felicella, Ramm Hering, Elizabeth Holliday, Cheyenne Johnson, Perry Kendall, Laura Knebel, Mona Kwong, Garth Mullins, Daniel Pare, Gerrard Prigmore, Samantha Robinson, Josey Ross, Andy Ryan, Aida Sadr, Christy Sutherland, Meaghan Umath, David Tu, Sharon Vipler, Je-West, Evan Wood, Steven Yau (2020). Risk Mitigation: In the Context of Dual Public Health Emergencies. Interim Clinical Guidance. British Columbia Centre on Substance Use. Available from: https://www.bccsu.ca/wp-content/uploads/2020/04/Risk-Mitigation-in-the-Context-of-Dual-Public-Health-Emergencies-v1.5.pdf

Public Health Agency of Canada guidance on vulnerable populations and COVID-19. Available from: https://www.canada.ca/content/dam/phac-aspc/documents/services/publications/diseases-conditions/coronavirus/covid-19-vulnerable-populations/vulnerable-populations-covid-19-eng.pdf

Pathway: Covid19 response. Hotel’s summary health needs assessment, May 2020. Brief Health Needs Assessment GLA COVID “Prevent” Limehouse and City hotels. Available from: https://www.pathway.org.uk/wp-content/uploads/Brief-HNA-Limehouse-and-City-22-May-20.pdf

Pathway: Clinical homeless sector plan: triage – assess – cohort – care (14 April 2020). Available from: https://www.pathway.org.uk/wp-content/uploads/COVID-19-Clinical-homeless-sector-plan-160420-1.pdf

COVID-19: guidance for commissioners and providers of hostel services for people experiencing homelessness and rough sleeping (2020). Public Health England. Available from: https://www.gov.uk/government/publications/covid-19-guidance-on-services-for-people-experiencing-rough-sleeping/covid-19-guidance-for-commissioners-and-providers-of-hostel-services-for-people-experiencing-homelessness-and-rough-sleeping#other-sources-of-information

Homeless link has also a collection of resources available for download on their website intended to helping services in the homelessness sector respond during the pandemic. Available here: https://www.homeless.org.uk/covid19-homelessness

## References

1 Ayano G, Shumet S, Tesfaw G, Tsegay L. A systematic review and meta-analysis of the prevalence of bipolar disorder among homeless people. BMC Public Health 2020; 20:731.

2 Ayano G, Solomon M, Tsegay L, Yohannes K, Abraha M. A Systematic Review and Meta-Analysis of the Prevalence of Post-Traumatic Stress Disorder among Homeless People. Psychiatr Q 2020; 91:949–63.

3 Beijer U, Wolf A, Fazel S. Prevalence of tuberculosis, hepatitis C virus, and HIV in homeless people: a systematic review and meta-analysis. Lancet Infect Dis 2012; 12:859–70.

4 Lewer D, Aldridge RW, Menezes D, et al. Health-related quality of life and prevalence of six chronic diseases in homeless and housed people: A cross-sectional study in London and Birmingham, England. BMJ Open 2019; 9. DOI:10.1136/bmjopen-2018-025192.

5 FEANTSA. European Federation of National Organisations Working with the Homeless. ETHOS - European Typology on Homelessness and housing exclusion. Brussels, 2005.

6 Amore K, Baker M, Howden-Chapman P. The ETHOS Definition and Classification of Homelessness: An Analysis. Eur J Homelessness 2011; 5:19–37.

7 Tsai J, Wilson M. COVID-19: a potential public health problem for homeless populations. Lancet Public Heal 2020; 5:e186–7.

8 Maxmen A. Coronavirus is spreading under the radar in US homeless shelters.Nature 2020; 581:129–30.

9 Conway B, Truong D, Wuerth K. COVID-19 in homeless populations: unique challenges and opportunities. Future Virol 2020; 15:331–4.

10 Mohsenpour A, Bozorgmehr K, Rohleder S, Stratil J, Costa D. Homelessness and COVID-19: a rapid systematic review. PROSPERO 2020 CRD420201807033. 2020. https://www.crd.york.ac.uk/prospero/display_record.php?ID=CRD42020187033

11 Jackson N, Waters E. Criteria for the systematic review of health promotion and public health interventions.Health Promot. Int. 2005; 20:367–74.

12 Egger M, Smith GD, Schneider M, Minder C. Bias in meta-analysis detected by a simple, graphical test. BMJ 1997; 315:629–34.

13 Sterne JA., Egger M. Funnel plots for detecting bias in meta-analysis. J Clin Epidemiol 2001; 54:1046–55.

14 StataCorp. Stata Statistical Software: Release 16. 2019.

15 Nyaga VN, Arbyn M, Aerts M. Metaprop: a Stata command to perform meta-analysis of binomial data. Arch Public Heal 2014; 72:39.

16 Sterne J, Harbord R, White I. An overview of meta-analysis in Stata. United Kingdom Stata Users’ Group Meetings 2010 11, Stata Users Group. 2010.

17 Moher D, Liberati A, Tetzlaff J, Altman DG. Preferred Reporting Items for Systematic Reviews and Meta-Analyses: The PRISMA Statement. PLoS Med 2009; 6:e1000097.

18 Mosites E, Parker EM, Clarke KEN, et al. Assessment of SARS-CoV-2 Infection Prevalence in Homeless Shelters — Four U.S. Cities, March 27–April 15, 2020. MMWR Morb Mortal Wkly Rep 2020; 69:521–2.

19 Baggett TP, Keyes H, Sporn N, Gaeta JM. Prevalence of SARS-CoV-2 Infection in Residents of a Large Homeless Shelter in Boston. JAMA 2020; 323:2191.

20 Marquez H, Ramers C, Northrup A, et al. Response to the COVID-19 pandemic among people experiencing homelessness in congregant living settings in San Diego, CA. Clin Infect Dis 2020; published online Oct 28. DOI:10.1093/cid/ciaa1668.

21 Yoon JC, Montgomery MP, Buff AM, et al. Coronavirus Disease 2019 (COVID-19) Prevalences Among People Experiencing Homelessness and Homelessness Service Staff During Early Community Transmission in Atlanta, Georgia, April– May 2020. Clin Infect Dis 2020; published online Sept 8. DOI:10.1093/cid/ciaa1340.

22 Imbert E, Kinley PM, Scarborough A, et al. Coronavirus Disease 2019 Outbreak in a San Francisco Homeless Shelter. Clin Infect Dis 2020; published online Aug 3. DOI:10.1093/cid/ciaa1071.

23 Tobolowsky FA, Gonzales E, Self JL, et al. COVID-19 Outbreak Among Three Affiliated Homeless Service Sites — King County, Washington, 2020. MMWR Morb Mortal Wkly Rep 2020; 69:523–6.

24 Henwood BF, Redline B, Lahey J. Surveying Tenants of Permanent Supportive Housing in Skid Row about COVID-19. J Health Care Poor Underserved 2020; 31:1587–94.

25 Samuels EA, Karb R, Vanjani R, Trimbur MC, Napoli A. Congregate Shelter Characteristics and Prevalence of Asymptomatic SARS-CoV-2. medRxiv 2020; : 2020.05.21.20108985.

26 O’Shea T, Bodkin C, Mokashi V, et al. Pandemic Planning in Homeless Shelters: A Pilot Study of a Coronavirus Disease 2019 (COVID-19) Testing and Support Program to Mitigate the Risk of COVID-19 Outbreaks in Congregate Settings. Clin Infect Dis 2020; published online June 8. DOI:10.1093/cid/ciaa743.

27 Ly TDA, Hoang VT, Goumballa N, et al. Screening of SARS-CoV-2 among homeless people, asylum seekers and other people living in precarious conditions in Marseille, France, March April 2020. medRxiv 2020; : 2020.05.05.20091934.

28 Lewer D, Braithwaite I, Bullock M, et al. COVID-19 among people experiencing homelessness in England: a modelling study. Lancet Respir Med 2020; 8:1181– 91.

29 Culhane D, Treglia D, Steif K, Kuhn R, Byrne T. Estimated Emergency and Observational/Quarantine Capacity Need for the US Homeless Population Related to COVID-19 Exposure by County; Projected Hospitalizations, Intensive Care Units and Mortality. 2020 https://escholarship.org/uc/item/9g0992bm.

30 Ramaswamy M, Hemberg J, Faust A, et al. Criminal Justice–Involved Women Navigate COVID-19: Notes From the Field. Heal Educ Behav 2020; 47:544–8.

## References

1 Tobolowsky FA, Gonzales E, Self JL, et al. COVID-19 Outbreak Among Three Affiliated Homeless Service Sites — King County, Washington, 2020. MMWR Morb Mortal Wkly Rep 2020; 69:523–6.

2 Yoon JC, Montgomery MP, Buff AM, et al. Coronavirus Disease 2019 (COVID-19) Prevalences Among People Experiencing Homelessness and Homelessness Service Staff During Early Community Transmission in Atlanta, Georgia, April–May 2020. Clin Infect Dis 2020; published online Sept 8. DOI:10.1093/cid/ciaa1340.

3 Ramaswamy M, Hemberg J, Faust A, et al. Criminal Justice–Involved Women Navigate COVID-19: Notes From the Field. Heal Educ Behav 2020; 47:544–8.

4 Culhane D, Treglia D, Steif K, Kuhn R, Byrne T. Estimated Emergency and Observational/Quarantine Capacity Need for the US Homeless Population Related to COVID-19 Exposure by County; Projected Hospitalizations, Intensive Care Units and Mortality. 2020 https://escholarship.org/uc/item/9g0992bm.

5 Lewer D, Braithwaite I, Bullock M, et al. COVID-19 among people experiencing homelessness in England: a modelling study. Lancet Respir Med 2020; 8:1181–91.

